# Adolescents with non-suicidal self-injury exhibit increased pain empathic neural reactivity and personal distress to physical but not affective pain

**DOI:** 10.1101/2025.09.10.25335218

**Authors:** Heng Jiang, Jingxian He, Bo Zhou, Yuxuan Guo, Xianyang Gan, Xiaoxia Fan, Xia Wang, Stefania Ferraro, Deniz Vatansever, Keith M Kendrick, Liyuan Li, Benjamin Becker

## Abstract

**Background:** Non-suicidal self-injury (NSSI) in adolescents represents a critical public health issue. While symptomatic links between NSSI and alterations in pain and social processing have been established, changes in neural responses and everyday reactivity to others’ pain remain unknown.

**Methods:** This pre-registered study examined pain empathic processing in unmedicated adolescents with NSSI (n=29) and healthy controls (n=33) using functional magnetic resonance imaging (fMRI). A validated paradigm assessed neural responses to physical pain versus affective pain observation and was combined with both univariate and machine learning analytic approaches.

**Results:** NSSI participants exhibited significantly increased neural reactivity during physical pain empathy in lateral prefrontal, insular, temporal, and the somatomotor network regions (all p<0.05, FDR-corrected), while affective pain processing remained intact. Machine learning analysis revealed distinguishable whole-brain signatures, with a physical pain empathic pattern achieving superior discrimination in NSSI. NSSI participants reported elevated personal distress to others’ negative experiences in everyday life, which was associated with enhanced limbic reactivity during physical pain empathy.

**Conclusions:** Findings identify domain-specific neural hyperreactivity to others’ physical pain in NSSI adolescents and elevated personal distress in daily life. These characteristics may represent predisposing alterations that facilitate engagement in self-harm or consequences of repeated engagement in NSSI that impact everyday social behavior.

## Introduction

Non-suicidal self-injury (NSSI), defined as the deliberate infliction of harm to one’s own body without suicidal intent (1–3), has been included in the DSM-5 as a psychiatric condition warranting further investigation (American Psychiatric Association, 2013).

The condition primarily affects adolescents and has emerged as a pressing public health issue with substantial implications for mental and physical health (3–6). Epidemiological studies report alarmingly high prevalence rates of NSSI in adolescents, with global 12-month prevalence rates in this age group around 22%, and lifetime prevalence rates ranging between 20-50% (4,7,8). While engagement in NSSI decreases from the early twenties, NSSI during adolescence represents a strong marker for high risk of psychiatric conditions in adolescence and during adulthood (9).

Emotion-related dysregulations and interpersonal difficulties have been increasingly recognized as core socio-affective features that contribute to the development and maintenance of NSSI (1,2,4,10). Individuals with a history of NSSI often exhibit heightened emotional reactivity and hyperarousal in the context of impaired emotion regulation abilities (11,12). Social functions have received less attention in NSSI, but initial studies suggest increased self-consciousness and increased vulnerability to interpersonal social stress, which may promote both long-term risk for NSSI and the acute engagement in NSSI behavior (13,14). Within an integrated model of NSSI, intrapersonal and interpersonal factors, such as impaired emotion regulation and enhanced interpersonal distress reactivity, are considered to interact with distal risk factors (e.g., genetic predisposition, childhood adversity) to render young individuals at an increased propensity to engage in NSSI and maintain NSSI as a progressively dysfunctional strategy to regulate aversive social-affective states (10).

Initial neuroimaging research has begun to determine the underlying neural basis. Within these studies, alterations during affective and pain-related processes have been reported, including increased neurofunctional reactivity towards negative emotional stimuli (15) and reduced insula reactivity towards physical pain yet greater reactivity towards self-administered cold stimulation (16,17). With respect to interpersonal processing, an initial study demonstrated a negative bias in social interaction in NSSI adolescents, underpinned by altered dorsomedial prefrontal cortex (dmPFC) and subgenual anterior cingulate cortex (sgACC) recruitment (18).

Empathy, including understanding and affective sharing of the pain experienced by others (pain empathy) (19,20), plays a crucial role in intrapersonal and interpersonal domains (21), including initiation and maintenance of successful social interaction (22). Altered pain empathic processing has been increasingly reported in multiple psychiatric disorders and has been related to disorder-related traits (e.g., He et al., 2025; Li et al., 2019).

Patients with major depressive disorder (MDD) reported lower pain empathic experience when watching painful videos in the context of decreased brain activation in the right somatosensory-related regions and left middle cingulate cortex, whereas greater activations in their left inferior frontal gyrus (25), with a more recent study indicating that the neurofunctional alterations in pain empathic reactivity vary as a function of the stimuli presented such that patients with depression – but not anxiety disorders – exhibited increased insular reactivity to facial expressions of pain (empathy for effective pain) (26), while patients suffering from migraine specifically exhibit increased insular and lateral prefrontal reactivity towards physical pain infliction in others (Liu et al., 2025).

Together, while neurofunctional pain empathy alterations have been increasingly reported in disorders related to socio-affective and pain dysregulations (23,24,27), alterations in adolescents with NSSI have not been examined. The rationale is further strengthened by recent reviews indicating that NSSI is associated with altered neural activation in core systems underlying pain, interpersonal, and self-related processing (6,28). Moreover, pain empathy and the underlying neural systems undergo a progressive development during adolescence (29–31) and, as such, may represent a sensitive marker for socio-affective development during this period.

In the current study, we applied a validated functional magnetic resonance imaging (fMRI) pain empathy paradigm (26,27,32,33) with univariate and machine-learning-based analysis methods to compare NSSI adolescents with healthy controls (HC), to investigate whether NSSI adolescents demonstrate altered pain empathic processing. Based on increasing evidence for domain-specific pain empathic dysfunctions (26,27), we examined reactivity towards painful facial expression or noxious stimulation of body limbs (33,34), respectively. Adolescents with NSSI have been suggested have deficits with the ability of facial expression recognition (35) and self-harm experiences may cause them to resonate more strongly with similar physical pain, we hypothesize that compared to HC, NSSI adolescents would exhibit increased empathic reactivity to physical pain in salience and arousal-related brain regions but decreased empathic reactivity to affective pain (Clinical trial preregistration see: ClinicalTrials.gov ID: NCT05968313).

## Methods

### Participants

Thirty adolescents with NSSI were recruited from a local hospital, and thirty-five healthy controls were recruited through advertisements. The general exclusion criteria for both groups included left-handedness, color vision deficiency, smoking, and a current or history of a neurodevelopmental (e.g., attention-deficit hyperactivity disorder, ADHD), neurological disorder, or contraindications to MRI. NSSI participants were diagnosed and screened according to DSM-5 in a clinical interview by an experienced clinician, with diagnosis being confirmed by a previously validated and sensitive self-report questionnaire (36). Additional inclusion criteria for the NSSI group include (1) meeting the diagnostic criteria of NSSI, (2) NSSI as the primary diagnosis and reason to seek treatment, and (3) no previous behavioral and pharmacological intervention for a psychiatric condition, including NSSI. The additional inclusion criteria of the HC group included no current or history of a psychiatric condition, including NSSI. Twenty-nine NSSI and thirty-three HC participants were included in the final analysis; two healthy participants were excluded due to excessive head motion (> 6 mm or 6°), and one NSSI participant did not complete the assessments.

Participants’ depression levels were assessed using the Patient Health Questionnaire (PHQ-9) (37), and the Interpersonal Reactivity Index (IRI) was used to measure the self-reported empathy (38,39). Based on validated self-report measures, the NSSI group exhibited significantly higher levels of depression and empathic interpersonal distress compared to the HC (Table 1).

**Table 1.**
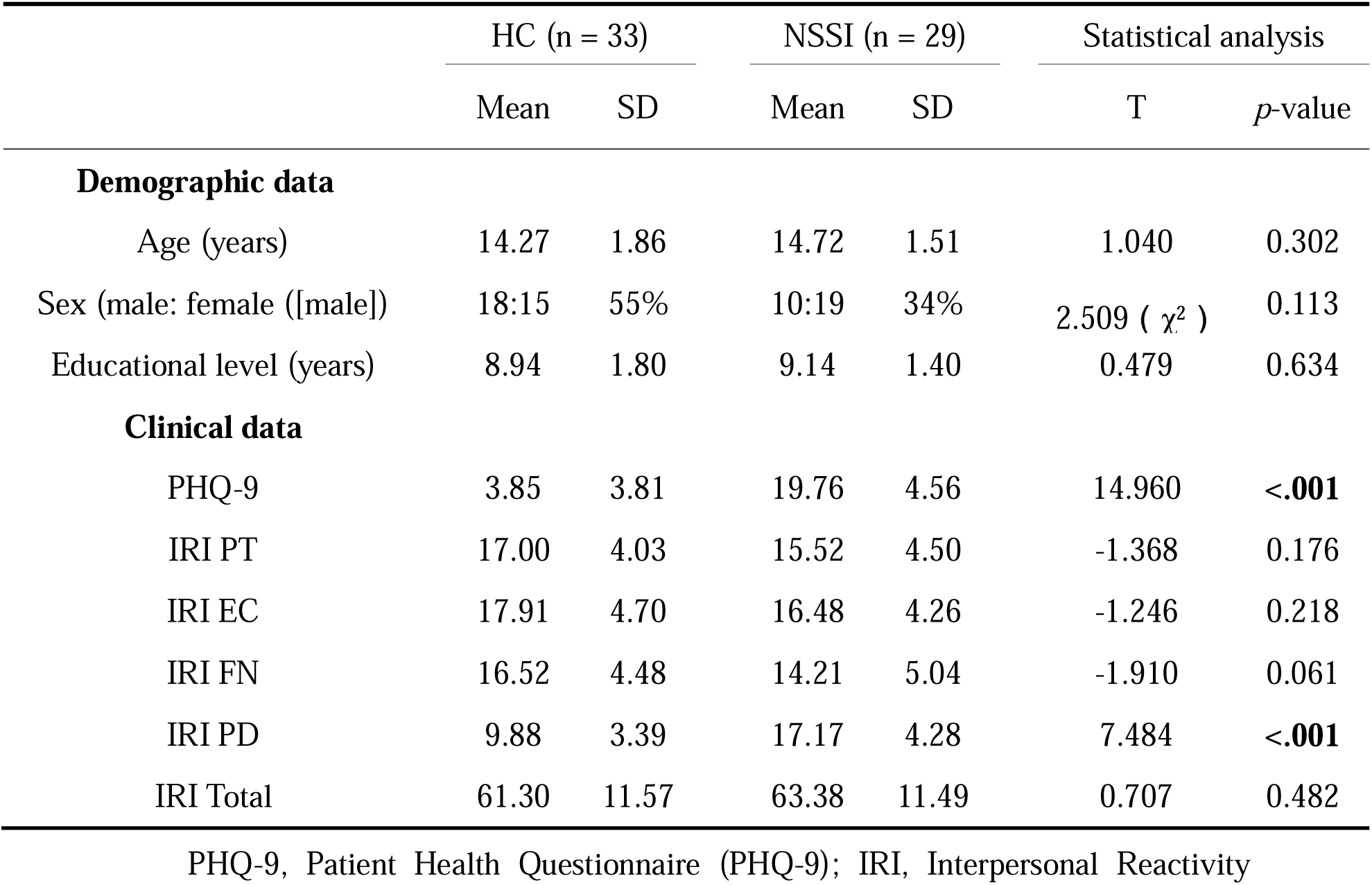
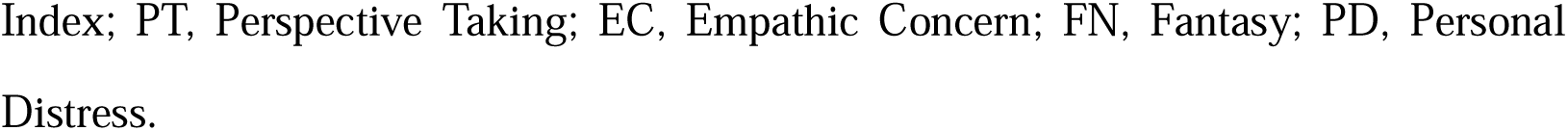
Demographics and clinical characteristics of HC and NSSI participants.

|  | HC (n = 33) |  | NSSI (n = 29) |  | Statistical analysis |  |
| --- | --- | --- | --- | --- | --- | --- |
|  | Mean | SD | Mean | SD | T | *p*-value |
| **Demographic data** |  |  |  |  |  |  |
| Age (years) | 14.27 | 1.86 | 14.72 | 1.51 | 1.040 | 0.302 |
| Sex (male: female ([male])) | 18:15 | 55% | 10:19 | 34% | 2.509 ( $\chi^2$ ) | 0.113 |
| Educational level (years) | 8.94 | 1.80 | 9.14 | 1.40 | 0.479 | 0.634 |
| **Clinical data** |  |  |  |  |  |  |
| PHQ-9 | 3.85 | 3.81 | 19.76 | 4.56 | 14.960 | **<.001** |
| IRI PT | 17.00 | 4.03 | 15.52 | 4.50 | -1.368 | 0.176 |
| IRI EC | 17.91 | 4.70 | 16.48 | 4.26 | -1.246 | 0.218 |
| IRI FN | 16.52 | 4.48 | 14.21 | 5.04 | -1.910 | 0.061 |
| IRI PD | 9.88 | 3.39 | 17.17 | 4.28 | 7.484 | **<.001** |
| IRI Total | 61.30 | 11.57 | 63.38 | 11.49 | 0.707 | 0.482 |

PHQ-9, Patient Health Questionnaire (PHQ-9); IRI, Interpersonal Reactivity
Index; PT, Perspective Taking; EC, Empathic Concern; FN, Fantasy; PD, Personal Distress.

All participants, and their legal guardians if under 18 years of age, provided written informed consent. All participants were compensated 100 RMB for participation. Protocols were pre-registered (ClinicalTrials.gov ID: NCT05968313), in accordance with the Declaration of Helsinki, and approved by the ethics committees of the University of Electronic Science and Technology of China and Sichuan Academy of Medical Sciences & Sichuan Provincial People’s Hospital.

### Stimuli and Paradigm

A validated fMRI paradigm was utilized to determine the neural bases of pain empathic processing (26,27,32,33), see Fig.1. Briefly, the paradigm presented four conditions, i.e. physical pain (PP), physical control (PC), affective pain (AP), and affective control (AC), by showing stimuli depicting noxious infliction of pain to the limbs in everyday situation from a first-person perspective (PP), matched control stimuli (PC) (40), stimuli depicting painful (AP), or matched neutral facial (AC) expression. The current paradigm employed a block design, consisting of 16 blocks in total. Each block contained four trials of the same condition. Every trial began with a white fixation cross (1 s), followed by the stimulus presentation (3 s). Between blocks, a red fixation cross was presented to allow participants to rest (8–12 s). To avoid cognitive modulation of empathic reactivity, participants were asked to passively view the stimuli (26,27,32,33).

**Fig 1.**
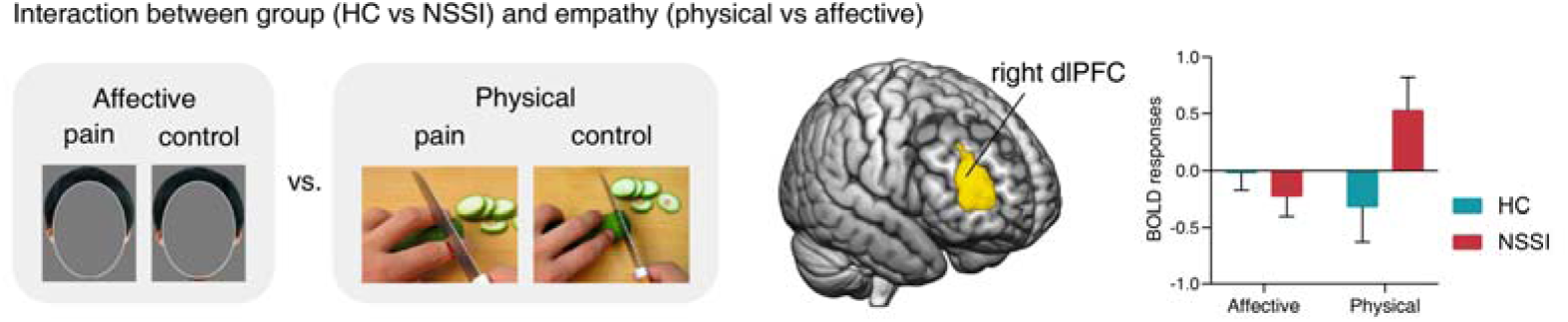
Significant interaction of group (HC vs. NSSI) and (empathy of affective vs. physical pain) located in the right lateral frontal cortex. Left, the significant cluster of the higher activation of HC participants compared to NSSI participants, during compare empathy of affective pain to physical pain. Clusters were formed at voxel-level *p*<0.001 and thresholded at *p*<0.05, cluster-level FDR-corrected. Right the mean BOLD signal of the significant cluster of the two groups during each contrast (affective: empathy of affective pian vs. affective control condition, physical: empathy of physical pian vs. physical control condition). To avoid potential copyright infringement, the original figure design was adapted, see (26,27,32,33) for details.

### MRI data acquisition and preprocessing

MRI data were collected on a 3.0 T scanner (Vida; Siemens Healthineers, Erlangen, Germany) with a 64-channel head coil. Structural images were acquired using high-resolution T1-weighted images by the original 3D Single-shot TurboFLASH sequence from Siemens (TR=2300 ms, TE=2.32 ms, FOV=240 mm, FA=8°, 256×256 matrix, 0.9×0.9×0.9 mm voxels, 192 sagittal slices, phase encoding posterior » anterior). Structural images were used for anatomical localization and warping to the standard Montreal Neurological Institute (MNI) space. Functional images were acquired with an echo planar imaging-free induction decay (EPI-FID) sequence (TR=2000 ms, TE=29 ms, FOV=240 mm, FA=90°, 80×80 matrix, 3×3×3 mm voxels, 36 interleaved ascending axial slices, phase encoding posterior » anterior).

MRI data were preprocessed using the Statistical Parametric Mapping (SPM 12, https://www.fil.ion.ucl.ac.uk/spm) toolbox. The first six volumes of fMRI images were removed, the different acquisition timing of each slice was corrected and realigned to the first volume, and nonlinear distortions related to the head motion were corrected by unwarping. The high-resolution anatomical image was segmented and co-registered with the functional images to generate the skull-stripped structural image. The functional images were normalized to MNI space, interpolated to 2×2×2 mm^3^ voxel size, smoothed by an 8-mm full-width at half maximum (FWHM) Gaussian kernel, and bias field corrected. Image intensity outliers were identified based on meeting any of the following criteria: (a) signal intensity >3 standard deviations from the global mean, (b) signal intensity and Mahalanobis distances >10 mean absolute deviations based on moving averages with a full-width at half maximum (FWHM) of 20 image kernels. The time points marked as outliers were severed as separate nuisance covariates included in the first-level model.

### Univariate analyses

Subject-level general linear model (GLM) analysis was conducted using SPM12. Twenty-four head motion parameters and indicator vectors of outlier time points were modeled as nuisance regressors (41–43). The fixation-cross epochs served as the implicit baseline, and the picture presentation period was used as the regressor of interest. A high-pass filter of 180s was applied.

We initially conducted a whole-brain voxel-wise ANOVA model with the factors group (NSSI vs HC) and pain empathic condition (physical vs. affective empathy, PP-PC vs. AP-AC) using a partitioned error approach. A significant interaction was followed by whole-brain voxel-wise two-sample tests on the respective pain empathic contrasts (PP vs. PC, AP vs. AC). Parameter estimates were extracted from significant clusters to disentangle interaction effects using MarsBar (http://marsbar.sourceforge.net/, Brett et al., 2002).

Given that the level of personal distress (PD), a subscale of the IRI referring to self-referenced negative arousal to others’ distress (38,39), has been linked to behavioral and neural pain reactivity (32,45,46), and has significant group differences in the current study, we examined associations between PD and brain activation using a multiple regression analysis in SPM.

The resulting statistical maps were threshold at a whole-brain level using a cluster-level FDR correction of *p*<0.05_FDR_ combined with a cluster-forming threshold of *p*<0.001_uncorrected_.

Considering the significantly higher depression level of NSSI adolescents, we conducted additional linear regression analysis for the NSSI group, with PHQ-9 rating as the independent variable and the parameter estimated from each significant cluster as the dependent variable.

### Machine-learning-based analysis

To determine whole-brain predictive accuracy of the neural alterations, we applied a validated multivariate brain decoder developed by our previous study (33), the vicarious pain-predictive pattern, to the dataset of the current two groups, separately. Based on the univariate results, we focused on physical pain empathy mainly (PP vs. PC) and tested the predictive performance of the NS (noxious stimulation) vicarious pain-predictive pattern to distinguish between con files of PP vs. PC by a forced-choice classification method, from the HC and NSSI group, separately.

We further used a permutation test to investigate whether the pattern expression of the decoder to the HC and NSSI shows a significant group difference (27,42). In this approach, the H_0_ assumes that the HC and NSSI groups have no significant differences. In each permutation, individual-level data were randomly selected from the two groups, scaled (divided) by the Z-score normalization method to reduce the impact of potential outliers, and then a forced-choice classification was performed, with 10,000 permutations. Finally, we tested whether the observed classification accuracy of the original single group (e.g., NSSI) has a significant difference (*p*<0.05) with the null distribution generated by permutation.

## Results

### NSSI group shows pain empathy-specific neurofunctional alterations

A significant interaction effect between group and pain empathic condition was observed in a large cluster located in the right dorsolateral prefrontal cortex (dlPFC) (extended to anterior inferior frontal sulcus (IFSa), see Fig.1 and Table 2). Plotting the parameter estimates of the right dlPFC cluster indicated that the interaction effect was driven by increased neural reactivity in the NSSI group towards physical pain empathy, while the HC showed decreased activation during this condition (Fig. 1).

**Table 2.**
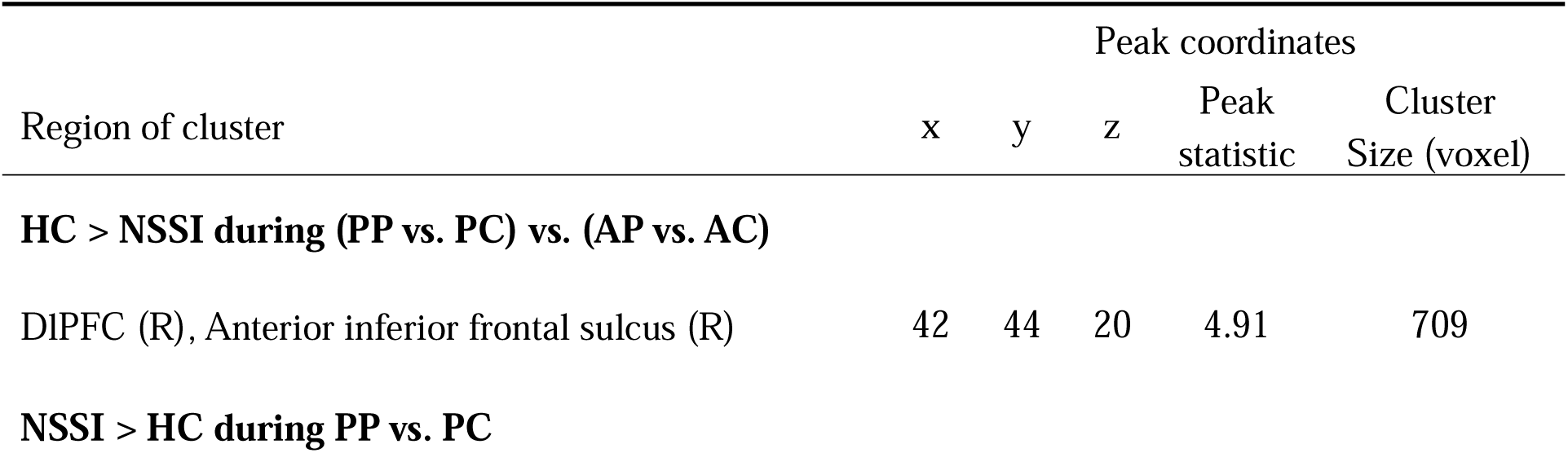
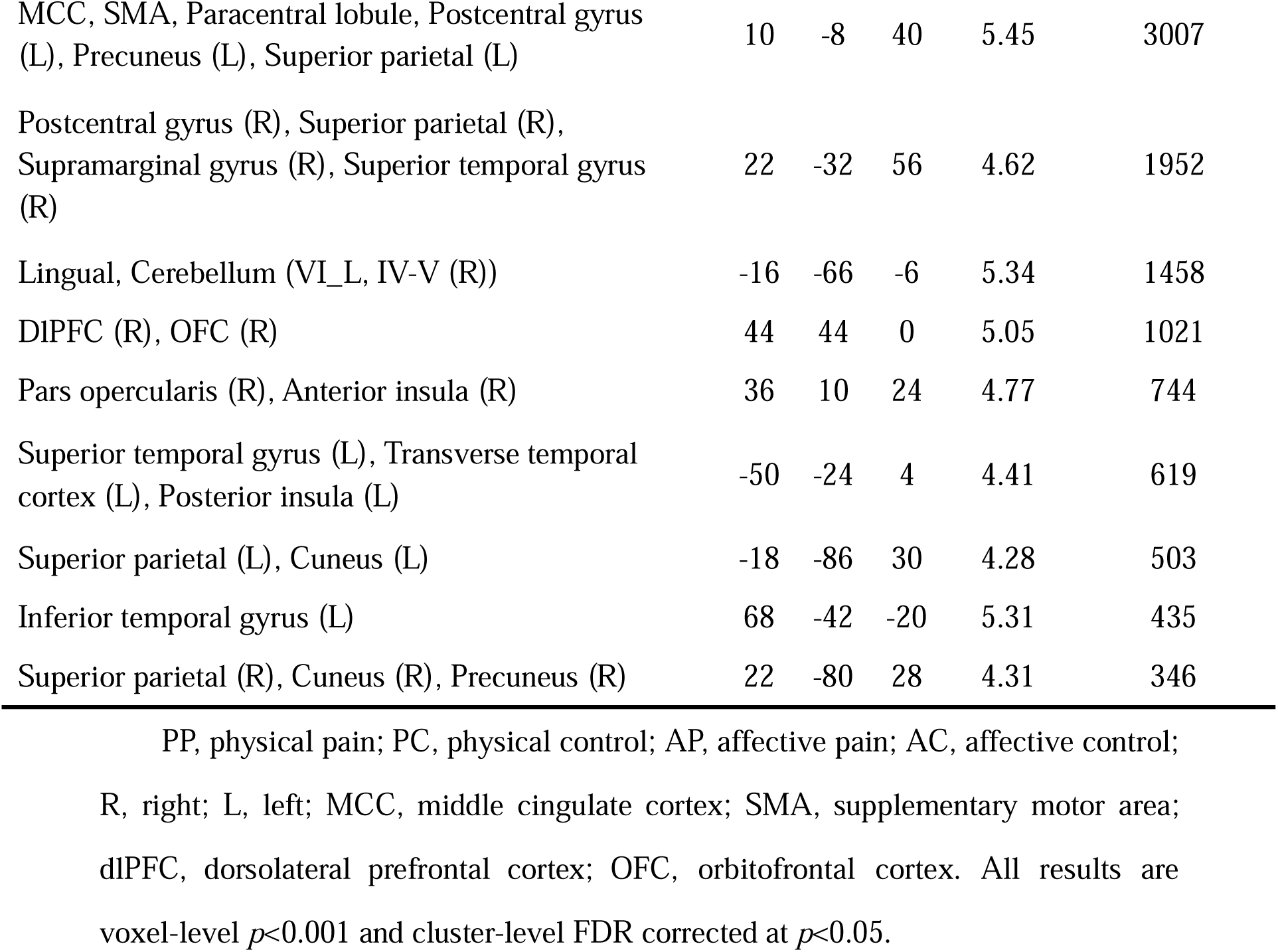
Regions show differential activations.

| Region of cluster | Peak coordinates |  |  |  |  |
| --- | --- | --- | --- | --- | --- |
|  | x | y | z | Peak statistic | Cluster Size (voxel) |
| **HC > NSSI during (PP vs. PC) vs. (AP vs. AC)** |  |  |  |  |  |
| DIPFC (R), Anterior inferior frontal sulcus (R) | 42 | 44 | 20 | 4.91 | 709 |
| **NSSI > HC during PP vs. PC** |  |  |  |  |  |
| MCC, SMA, Paracentral lobule, Postcentral gyrus (L), Precuneus (L), Superior parietal (L) | 10 | -8 | 40 | 5.45 | 3007 |
| Postcentral gyrus (R), Superior parietal (R), Supramarginal gyrus (R), Superior temporal gyrus (R) | 22 | -32 | 56 | 4.62 | 1952 |
| Lingual, Cerebellum (VI_L, IV-V (R)) | -16 | -66 | -6 | 5.34 | 1458 |
| dIPFC (R), OFC (R) | 44 | 44 | 0 | 5.05 | 1021 |
| Pars opercularis (R), Anterior insula (R) | 36 | 10 | 24 | 4.77 | 744 |
| Superior temporal gyrus (L), Transverse temporal cortex (L), Posterior insula (L) | -50 | -24 | 4 | 4.41 | 619 |
| Superior parietal (L), Cuneus (L) | -18 | -86 | 30 | 4.28 | 503 |
| Inferior temporal gyrus (L) | 68 | -42 | -20 | 5.31 | 435 |
| Superior parietal (R), Cuneus (R), Precuneus (R) | 22 | -80 | 28 | 4.31 | 346 |

PP, physical pain; PC, physical control; AP, affective pain; AC, affective control; R, right; L, left; MCC, middle cingulate cortex; SMA, supplementary motor area; dIPFC, dorsolateral prefrontal cortex; OFC, orbitofrontal cortex. All results are voxel-level $p < 0.001$ and cluster-level FDR corrected at $p < 0.05$ .

Condition-specific voxel-wise whole-brain two-sample t-tests directly comparing the groups revealed that the NSSI groups exhibited higher physical pain empathic reactivity (contrast PP vs. PC) in a widespread bilateral network encompassing the right dlPFC, anterior insula, and inferior frontal gyrus (pars opercularis) as well as bilateral middle cingulate cortex (MCC), postcentral gyrus, STG, lingual gyrus, supramarginal (inferior frontal gyrus), left posterior insula, and precuneus (see Fig.2 and Table 1). For affective pain, no significant group differences were observed.

**Fig 2.**
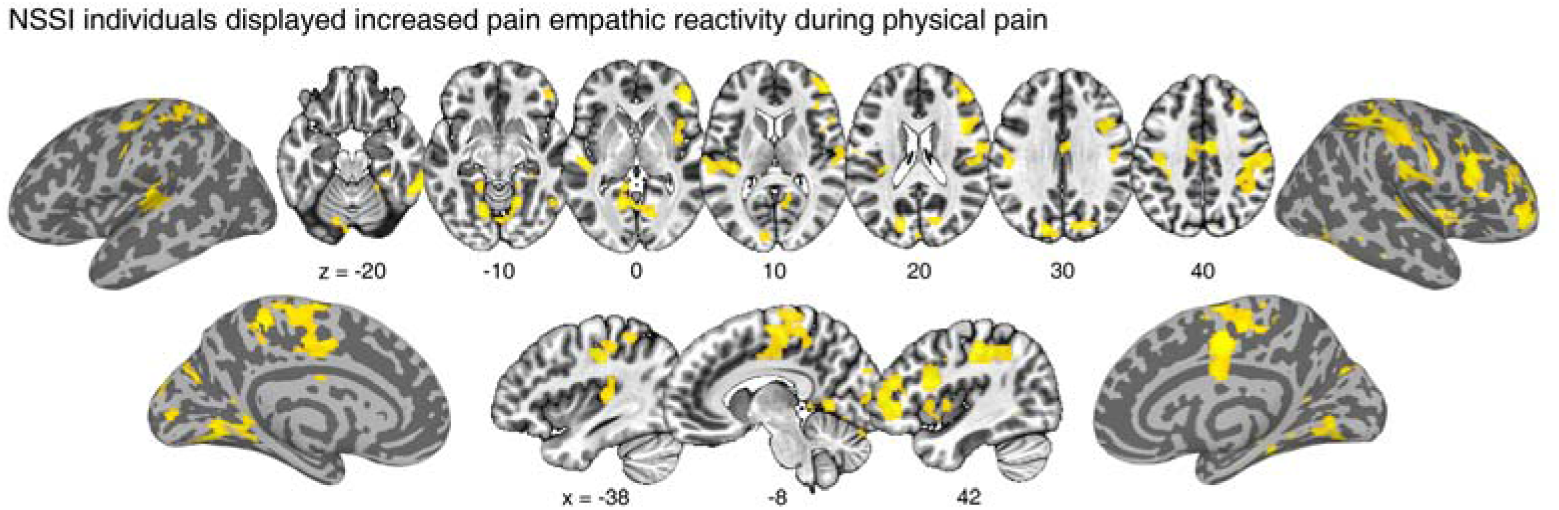
Higher brain activation of NSSI than HC participants during empathy of physical pain. Brain regions with higher activation of NSSI than HC participants during physical pain empathic reactivity displayed at the voxel-level *p*<0.001 and cluster-level FDR corrected at *p*<0.05.

### Associations with levels of interpersonal distress and depression

The results of the whole-brain voxel-wise regression analysis revealed that – in the entire sample – higher levels of PD, which has been linked to behavioral and neural pain reactivity (32,45,46), were significantly associated with physical pain empathic reactivity in a widespread network encompassing limbic and temporal regions involved in social and affective processing, including the amygdala, temporal pole and fusiform gyrus as well as lingual and anterior cerebellar regions (voxel-level threshold *p*<0.001 and cluster-level FDR corrected *p*<0.05, see Fig.3 and Table 3). No significant results were observed for the affective pain empathy.

**Fig 3.**
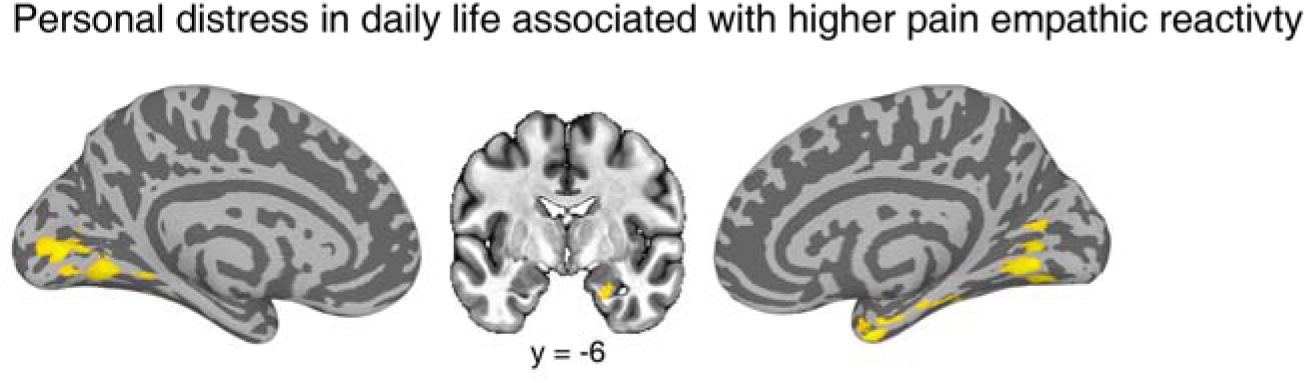
Regions showing increased activity as a function of higher personal distress during physical pain empathy. Regions with positive correlations were established between the self-reported PD rating and brain activation under the PP vs. PC condition. Voxel-level threshold *p*<0.001 and cluster-level FDR corrected at *p*<0.05.

**Table 3.** Regression analysis between personal distress and brain activation in all participants.

| Region of cluster | Peak coordinates |  |  | Peak statistic | Cluster Size (voxel) |
| --- | --- | --- | --- | --- | --- |
|  | x | y | z |  |  |
| Lingual, Cerebellum (VI, IV-V (R)) | 14 | -58 | -10 | 4.80 | 1643 |
| Amygdala (R), Temporal pole (R), Fusiform (R), Parahippocampal gyrus(R) | 22 | 6 | -6 | 4.34 | 597 |

Regression analysis between levels of depression and activity in the brain regions exhibiting altered activity in NSSI revealed that the independent variable (depression level) was a significant predictor of the dependent variable – empathy response of physical pain at the largest cluster (MCC, SMA, paracentral lobule, left postcentral gyrus, left precueus, left superior parietal; *t*=2.073, *p*=0.048, unstandardized coefficient=0.056). No significant associations were found between levels of depression and activation in other clusters, including the cluster exhibiting the significant interaction effect in the ANOVA (all *p*>0.05)

### NSSI group exhibits altered whole-brain expressions of pain empathic reactivity

The multivariate NS vicarious pain-predictive pattern significantly and accurately distinguished between PP vs. PC in both HC (accuracy 88% (±5.7% s.e.m.), Cohen’s d=1.72, sensitivity 88% (95% CI 77–98), specificity 88% (95% CI 75–97), two-sided binomial test *p* = 1.10×10) and NSSI group (accuracy 93% (±4.7% s.e.m.), Cohen’s d=1.79, sensitivity 93% (95% CI 83–100), specificity 93% (95% CI 82–100), *p*=2.00×10), see Fig.4. Further permutation analysis suggested that there are significant differences between the pattern expression of NSSI and HC participants at the whole brain level (*p*=1.10×10³).

**Fig 4.**
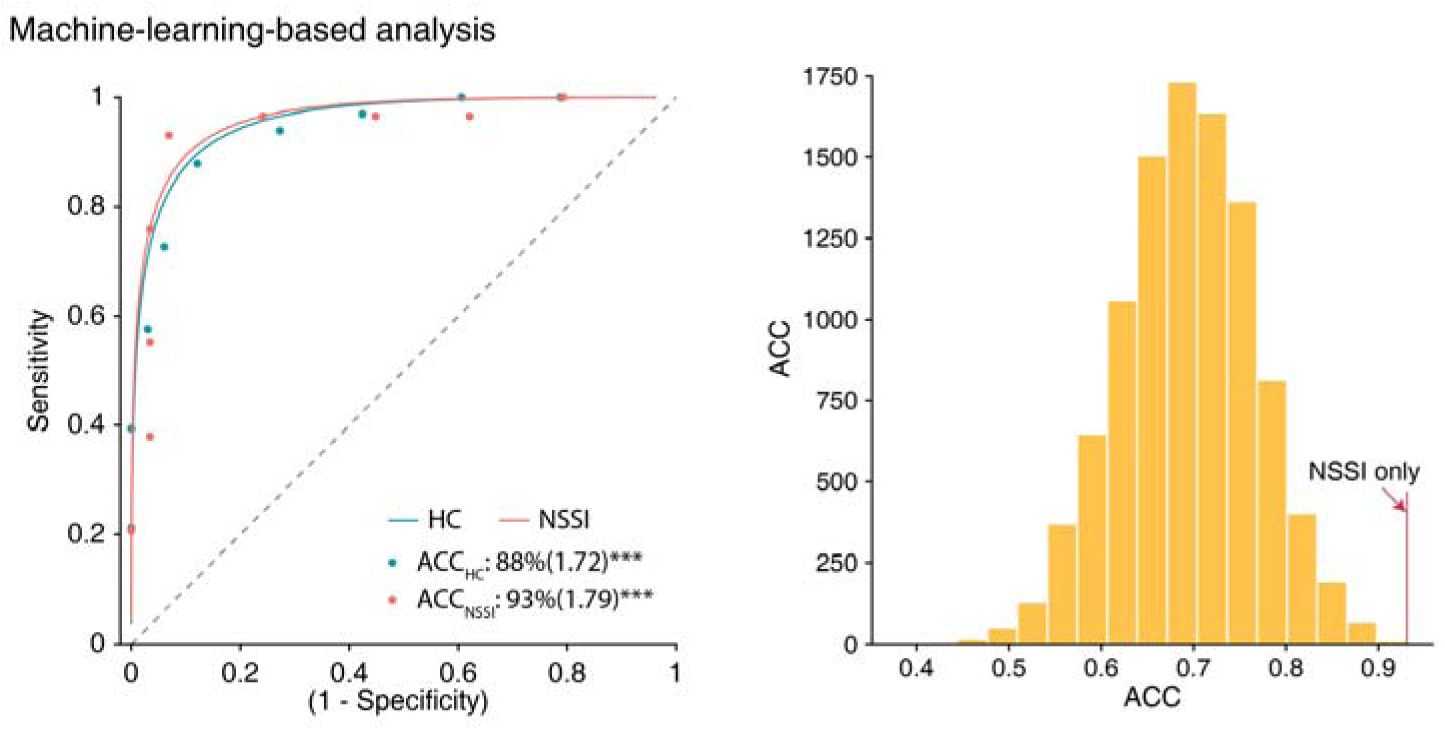
Test with empathy decoder of physical pain. The receiver operating characteristic (ROC) plot illustrates that the NS vicarious pain-predictive pattern could accurately detect pain empathic reactivity in terms of classifying physical pain vs. the physical control, in both HC (n=33) and NSSI (n=29) groups, separately (left). The premutation results indicated that the pattern expression of the two groups exhibits significant differences (right).

## Discussion

To our knowledge, this is the first study to explore the neurofunctional basis of pain empathic processing in adolescents with NSSI behavior. Combining fMRI with univariate and multivariate machine-learning-based analysis revealed that adolescents with NSSI behaviors exhibit increased neurofunctional pain empathic reactivity in a widespread network encompassing right lateral prefrontal regions (dLPFC, IFG), the insula, bilateral superior temporal regions, and core nodes of the somatomotor network (SMA, MCC, postcentral, and superior temporal gyrus) in response to others’ physical pain infliction, but intact processing of socio-affective (facial) pain signals. Multivariate analyses furthermore indicated that the physical pain predictive pattern can accurately predict pain empathic reactivity and can discriminate NSSI individuals from the HC group, underscoring the potential to serve as a neuromarker for altered pain empathic reactivity. On the behavioral level, NSSI individuals reported higher levels of PD, with the latter trait associated with higher neurofunctional reactivity in limbic, temporal, and visual regions. Follow-up analyses indicated that levels of depression were associated with pain empathic reactivity in the somatomotor network but not lateral prefrontal and insular regions. Together, the findings support our hypothesis that NSSI adolescents exhibit altered empathy processing, specifically to others’ physical pain, and highlight the contributing role that personal distress and depression may play in the observed neurofunctional alterations.

On the neural level the physical pain empathic stimuli elicited a stronger neural response in the NSSI individuals as compared to the HC in a widespread network encompassing regions commonly associated with experiencing one’ own and other people’s pain (e.g., postcentral gyrus, anterior insula; Fallon et al., 2020; Jauniaux et al., 2019b; Pavlova et al., 2021; Zhou et al., 2020), lateral prefrontal regions involved in the subjective experience of negative affect, including pain experience (e.g., Čeko, 2022; X. Liu et al., 2024; Zhang et al., 2025), the cortical mirror neuron system (e.g., pars opercularis of the inferior frontal gyrus and inferior parietal lobule) supporting pain empathy by imitation (53–57) and core regions of the somatomotor network, i.e., SMA, MCC, and postcentral gyrus, has been consistently identify as crucial network involved in empathy processing by meta-analysis review (58–60). Importantly, while the present study did not encompass subjective pain ratings, a neurofunctional decoder sensitive to the affective pain experience (33) showed a higher predictive accuracy in the NSSI group, suggesting a higher level of negative affect in the NSSI group.

Adolescents with NSSI behaviors may thus have a stronger aversive experience and resonance with pain inflicted in others, possibly (partly) because these stimuli resemble their own experience with noxious manipulation. In support of this, the NSSI individuals reported higher levels of PD, suggesting that NSSI adolescents have significantly stronger self-referenced negative arousal to others’ distress in everyday life. Regression analysis further indicated that higher PD is associated with higher pain empathic activity in limbic, temporal, and visual regions involved in social-affective processing (61), including the amygdala (62), temporal pole, and fusiform gyrus (34,63).

Contrary to our hypothesis, we did not observe alterations during affective pain processing induced by observing others’ facial expressions of pain. A similar pattern of altered neurofunctional reactivity in inferior frontal and insular regions during physical pain empathy has been reported in patients with migraine (27) as well as in individuals with autism spectrum disorder (64) and high levels of alexithymia (24), while patients with depression exhibited increased reactivity to affective pain in these regions (X. Xu et al., 2020, however see also Rütgen et al., 2021). One explanation of the physical pain empathy specific changes in NSSI may refer to a predisposing higher salience of physical pain in NSSI individuals, with previous studies supporting altered sensitivity to self-experienced pain in NSSI (66), while a shift in dispositional emotional – as compared to cognitive – empathy has been related to higher levels of emotion-induced NSSI in individuals with autism (67). A second explanation would align with the Perception-Action Model (PAM) of empathy, which proposes that past experience and similarity can influence pain empathic reactivity (68). The NSSI have undergone several instances of noxious self-infliction of pain to the limbs, and as such, this experience may have shaped their reactivity to observing similar physical pain in others. Consequently, the observed physical-empathy specific alterations may reflect predisposing dispositional features that render individuals vulnerable to developing NSSI or may reflect that repeated self-induced pain infliction leads to a higher sensitivity to similar pain infliction in others.

Pain empathy plays a crucial role both in intrapersonal and interpersonal domains (21,22), with both being closely related to NSSI behavior (1,2,4,10). Empathy may present a promising therapeutic target to reduce emotional reactivity and NSSI behavior, in particular given that traditional psychotherapeutic approaches (e.g., cognitive-behavioral therapy, Butler et al., 2006), non-invasive interventions (e.g., transcranial magnetic stimulation, Wu et al., 2025; neurofeedback, Y. Zhang et al., 2023), or even online training (see Yeager et al., 2022) show potential for helping adolescents with NSSI strengthen their capacity to modify empathy.

Findings need to be interpreted in the context of potential limitations inherent to clinical neuroimaging studies. First, the cross-sectional nature of the design does not allow us to disentangle predisposing alterations from those induced by repeated self-harm. Secondly, in line with previous studies reporting a high co-morbidity between NSSI and depression (18,72,73), NSSI individuals in the present study reported elevated levels of depression. Results from our regression analysis indicate that, in particular, alterations in the somatomotor network were related to depression, and future studies are required to disentangle interactions between depression and NSSI in adolescents.

Summarizing, the present study revealed enhanced neurofunctional pain reactivity in response to physical but not affective pain in adolescents with NSSI in the context of elevated PD in daily life. Brain functional alterations may represent a predisposition for or consequence of repeated NSSI behavior and a possible modifiable therapeutic target.

## Data Availability

Data availability is not allowed due to institutional and ethical restrictions.

## Acknowledgments

This work is supported by the National Natural Science Foundation of China (NSFC 82271583), the Ministry of Science and Technology of China (STI 2030–Major Projects 2022ZD0208500), the Hong Kong University Grants Council (GRF 17615525), the University of Hong Kong seed funding and start-up schemes (2407102536; 2402101713), and the Sichuan Science and Technology Program (2023YFS0023; 2024NSFSC1572).

BB, BZ, LYL obtained funding and guided the writing of articles with HJ, JXH, BZ, YXG, XXF, XW and LYL collected the data. HJ analyzed the data and drafted the manuscript. HJ and BB contributed to the interpretation of the results, critical revision of the manuscript, and approved the final version. LYL, XYG, SF, DV, KMK provide important suggestions and critical comments on the manuscript. All authors have read and approved the final manuscript.

